# What Do Clinicians Edit in Ambient AI-Drafted Clinical Documentation? A Qualitative Content Analysis

**DOI:** 10.64898/2026.01.05.26343473

**Authors:** Yawen Guo, Di Hu, Ziqi Yang, Seungjun Kim, Brian Tran, Jamie Lee, Sitha Vallabhaneni, Rachael Zehrung, Sairam Sutari, Steven Tam, Emilie Chow, Danielle Perret, Deepti Pandita, Kai Zheng

## Abstract

**Objective:** Ambient artificial intelligence (AI) documentation is increasingly used to draft clinical notes from patient-provider conversations, but how clinicians revise and finalize these drafts is not well understood. This qualitative content analysis study characterizes real-world edits to AI-generated drafts and identifies opportunities for improvement of AI design and the implementation process.

**Materials and Methods:** Eight coders analyzed clinical documentation generated by ambient AI from 200 clinical encounters. We developed an inductive coding framework with 11 codes across three categories: clinical content, terminology, and language style. Interrater reliability was assessed using Cohen’s kappa. We then applied thematic analysis to synthesize patterns across the coded edits.

**Results:** The most frequently edited content pertained to clinical facts including orders (e.g., procedures, lab tests) (40.0%), symptoms (30.3%), medication prescriptions (27.3%), and diagnosis descriptions (25.9%). In comparison, edits related to terminology use (11.6%) and language style (7.2%) were less frequent. The results of our thematic analysis show that most edits can be categorized into one of the following five types: to correct factual errors, to address needs of medical specialty, to express diagnostic certainties, to convert patient expressions into objective assessments recorded in medical terms, and to reorganize or condense content.

**Conclusion and Discussion:** Clinicians routinely revise ambient AI drafts to improve accuracy and clinical specificity. Future work on AI development and clinical implementation should emphasize specialty customization and support personalized documentation practices, alongside clinician education that promotes robust and consistent review routines to ensure documentation quality.

## Introduction

Ambient artificial intelligence (AI) systems are increasingly integrated into clinical workflows to support documentation by generating note drafts. [1,2] Early studies suggest they may reduce documentation burden, improve efficiency, and enhance note quality. [3,4] However, despite growing adoption, the real-world reliability and clinical appropriateness of AI-drafted documentation remain underexplored.

Prior evaluations have largely emphasized clinician and patient perspectives through surveys and interviews, focusing on usability, trust, and time savings rather than the content of the generated text. [5–9] A few studies compared AI-generated notes with reference text using automated performance and text-similarity metrics including the F1 score, overlap-based measures such as Recall-Oriented Understudy for Gisting Evaluation (ROUGE) and embedding-based scores. However, these approaches typically produce aggregate, note-level outcomes without explicating what content was changed. [10–12] Expert-based note quality assessments, such as structured reviewer ratings and instruments like the Physician Documentation Quality Instrument, provide an overall quality assessment but are often conducted on simulated or curated data, offering limited insight into the specific edits clinicians make or the reasons behind them. [13]

Limited work has compared real-world ambient AI drafts and final notes committed to electronic health record (EHR) systems to directly characterize editing behavior. Understanding these edits is essential because they show where AI drafts fall short and can reveal systematic gaps with implications for clinical safety, billing, and communication. [14] They can also inform model improvement and implementation support to better align ambient AI with clinical documentation practice.

To address this gap, we conducted a content analysis of 314 ambient AI-generated note sections from 200 paired ambient AI drafts and clinician-finalized versions, including 1,804 word-level edit operations (insertions, deletions, and replacements) across 33 specialties and 73 clinicians in an ambient AI pilot program. Using qualitative coding and thematic analysis, we examined the types of textual changes clinicians made, the content areas most frequently modified, and the broader themes that describe how clinicians refine AI-generated drafts into finalized documentation. This study offers a fine-grained empirical characterization of real-world clinician editing in ambient AI documentation, with the goal of informing more reliable and more workflow-aligned documentation support systems.

## Method

This study employed qualitative coding and thematic analysis to characterize clinician edits to AI-generated clinical note drafts at the content level. The research was conducted as part of a quality improvement initiative at the University of California, Irvine Medical Center (UCI Health), involving clinicians across ambulatory primary and specialty care clinics using two commercially available ambient AI systems that were integrated into the Epic electronic health record (EHR) via mobile and web interfaces. Vendors are referred to as Vendor A and Vendor B to preserve contractual and institutional confidentiality. The study protocol was approved by the University of California, Irvine Institutional Review Board (IRB #7123). All data processing occurred within HIPAA-compliant secure computing environments, including UCI Health’s Protected Virtual Computing Environment (PVCE) and Amazon Web Services (AWS) infrastructure.

### Data Source and Sampling Strategy

Ambient AI drafts clinical content by sections, covering history of present illness (HPI), assessment and plan (A&P), physical exam, and results, which clinicians can selectively insert into EHR notes. The overall corpus of this study consisted of all AI-generated draft note sections and their corresponding clinician-finalized versions recorded between September 26, 2024, and August 5, 2025, from the University of California, Irvine Medical Center. To construct an annotation dataset for content analysis that reflects the full corpus, we drew a stratified sample of 200 notes, each containing one or more AI-drafted note sections with clinician edits, representing 33 clinical specialties. Sampling quotas were aligned with the overall corpus distribution, ensuring the inclusion of at least two notes per specialty or one in cases where only a single note was available. Quotas were also designed to achieve a balanced representation from both vendor systems in the annotated sample.

### Edit Detection Using the Myers Diff Algorithm

We detected editing activity in the sample by quantifying insertions, deletions, and replacements between each ambient AI draft and the clinician-finalized note section. For each draft–final pair, we applied a word-level Myers diff algorithm to align tokens and identify edit operations, recording the span and sequence of each change. [15] We also generated side-by-side HTML views with highlighted edits, allowing for direct comparison of the AI output against clinician revisions. Figure 1 provides a synthetic example for illustration only and does not contain patient data.

**Figure 1.**
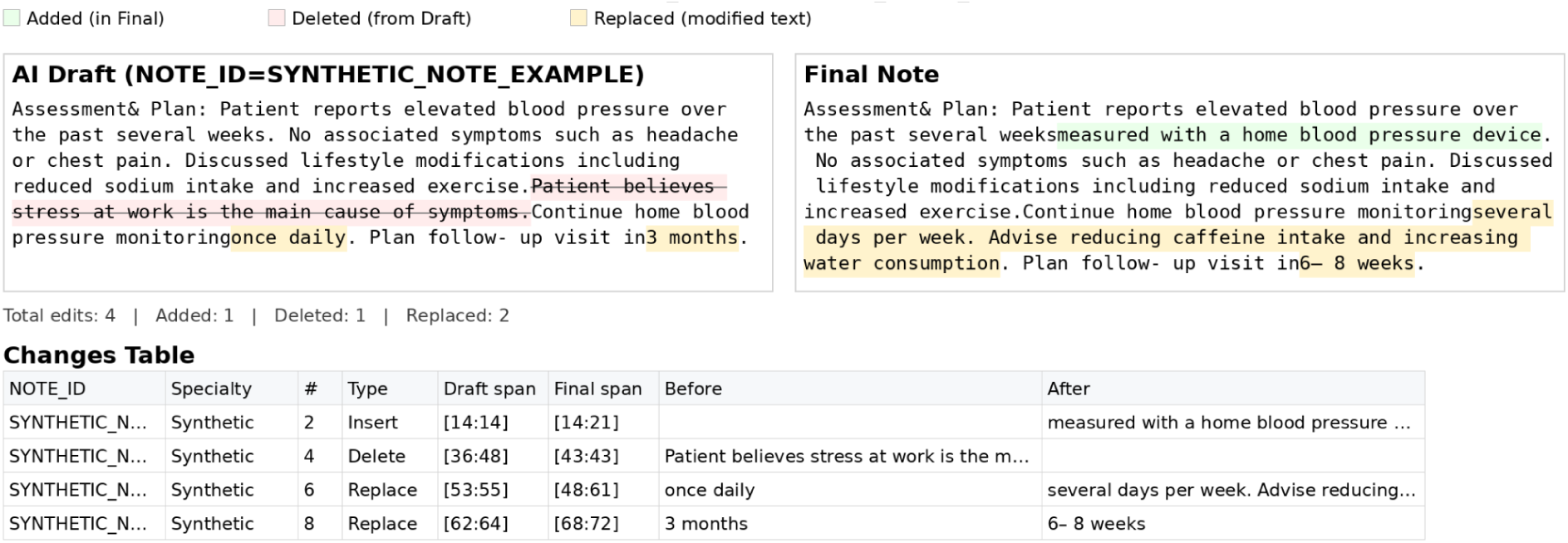
Synthetic Example of Side-by-Side Visualization of Clinician Edits to an AI-Generated Clinical Note.

### Sentence-Level Content Analysis

We defined an edit unit as the smallest contiguous set of sentences that fully captured each diff-detected edit span, including adjacent or overlapping edits, to preserve clinical context and meaning. To align draft and final text at the sentence level, we used a dynamic approach that allowed flexible mappings, including one-to-one, one-to-many (split), many-to-one (merge), and many-to-many alignments. Each edit unit was labeled by its alignment pattern and quantified by the number of insertions, deletions, and replacements it contained, capturing both within-sentence phrasing changes and larger structural revisions.

### Coding Framework and Procedure

An inductive coding framework was developed to characterize the types and functions of clinician edits. [16] Two researchers (YG, DH) independently reviewed a subset of edit units to develop an initial codebook. To support comprehensive inductive discovery across heterogeneous documentation patterns, we developed the codebook using the pooled set of note section pairs from 200 note spanning both vendor systems and a broad range of clinical specialties. The codebook was then iteratively refined through consensus meetings and multiple rounds of coding with eight coders, all with experiences in health informatics research and qualitative analysis. The full coding team then conducted an initial calibration round in which all eight coders applied the draft codebook to the same set of edit units; discrepancies were resolved through adjudication meetings and email discussion, and the codebook was iteratively refined. Interrater reliability was assessed using Cohen’s κ across major code categories. [17] We finalized the codebook after achieving acceptable interrater agreement across coders. The final refined framework consisted of three top-level code categories capturing distinct categories of editing behavior (entity-level clinical content changes, terminology and wording adjustments, or language and style modifications), along with 11 subcodes representing more granular phenomena. [18,19]

Using the finalized codebook, coders applied labels to each sentence-level edit unit. We treated each code as a binary label, present or absent, for each sentence-level edit unit, allowing for multiple codes per unit. Across eight coders, we computed Cohen’s kappa for each code for each annotator pair and macro-averaged kappas across codes with equal weight; overall reliability was the mean of the 28 pairwise macro-averaged kappa values. Because edit units often reflected specialty-specific documentation practices and patient-specific clinical context, we anticipated partial overlap at code boundaries and therefore used iterative calibration and consensus adjudication to ensure a stable codebook prior to thematic synthesis. All coding was conducted in structured spreadsheets using the finalized codebook. To maintain independent coding, each coder worked in a separate spreadsheet. For reference when needed, coders could consult color-coded HTML diff visualizations of the paired draft and final text to view each edit unit alongside the exact textual changes within the full note section.

### Thematic Analysis

After all edit units were coded using the finalized codebook, we conducted a thematic synthesis to identify higher-level patterns in clinician revision behavior. Vendor identities were masked during codebook development and edit-unit coding to reduce bias and support consistent application of the codebook. After coding was complete, vendors were specified for a vendor-stratified descriptive summary of code distributions and to assess whether themes were represented in both vendor groups. Two lead researchers (YG, DH) independently reviewed code distributions, representative edit units, and analytic memos to examine how frequently co-occurring codes reflected shared underlying revision behavioral patterns. Codes were iteratively grouped based on conceptual similarity and the clinical intent they represented (e.g., factual correction, clinical precision, documentation standardization). Through this process, we aggregated code-level edits into five higher-level themes that capture recurring clinician revision patterns in AI-drafted notes. Themes were refined through iterative team discussion and disagreements in theme boundaries were resolved through consensus. The resulting themes represent interpretive syntheses of multiple code types and are intended to characterize how clinicians systematically revise AI-drafted documentation.

## Results

### Dataset Characteristics

A total of 314 note section pairs from 200 notes containing ambient AI drafted sections and finalized versions were included in the annotated sample, representing 73 clinicians, 200 patients, and 33 clinical specialties. Clinicians could generate more than one ambient AI section draft within the same note (for example, only A&P, or both HPI and A&P). We coded section-level draft–final pairs, grouped by note ID for tracking. The complete sampled notes for qualitative annotation included a history of present illness section (n = 149) and an assessment and plan section (n = 135), while fewer included physical examination (n = 18) or results (n = 12). Across all sampled draft–final section pairs, clinicians made 1,804 token-level modifications, including 556 insertions, 393 deletions, and 855 replacements. The sample covered 33 specialties; family practice contributed the most notes (n = 54), followed by rheumatology (n = 22) and internal medicine (n = 20), with the remaining notes distributed across a broad set of lower-volume specialties (Table 2).

**Table 1.** Qualitative coding schema.

| Primary Code Category | Code | Example(s) | Description |
| --- | --- | --- | --- |
| Entity-level: Clinical Content | Medication (E-Med) | Before: “Keppra one pump BID”;<br>After: “Keppra 500 mg BID” | Edits that add, remove, or modify medication information, including medication name, dose, frequency, route, formulation, timing, or adherence details. |
|  | Symptom (E-Sym) | Before: “Cramping”;<br>After: “Cramping at night” | Edits that add, remove, or modify symptom mentions, including presence/absence, severity, location, timing, duration, triggers, or associated features. |
|  | Diagnosis (E-Dx) | Before: “epilepsy”;<br>After: “refractory focal epilepsy” | Edits that add, remove, or modify diagnoses/conditions or diagnostic labels, including specificity (type/subtype), qualifiers, or diagnostic framing. |
|  | Procedure / test / lab order (E-Test) | Before: “EEG not done”;<br>After: *(deleted)* | Edits to tests or procedures and related orders, including labs and imaging, whether performed in-clinic or externally, such as changes to test type, status, referrals, timing/location, or results. |
|  | Demographics / ID (E-ID) | Before: “[WRONG-SPELLED NAME]”;<br>After: “[CORRECTED NAME]” | Edits that add, remove, or modify patient identifiers or demographic details (e.g., age, name, pronouns, PHI placeholders) |
|  | Social context (E-Soc) | Before: *(none)*;<br>After: “lives with spouse” | Edits that add, remove, or modify social history/context (e.g., tobacco/alcohol/substance use, living situation, education, occupation, support system, marital status, habits/behaviors) [20] |
|  | Past medical history (E-Hx) | Before: (none);<br>After: “Previously saw nephrologist at [LOCATION]” | Edits that add, remove, or modify past medical history, including comorbidities, prior events, prior specialty care, surgeries, and relevant historical conditions. |
| Terminology and Wording | Lay–jargon substitution (T–Lay) | Before: “pain”;<br>After: “myalgia” | Edits that substitute clinical jargon with lay language (or vice versa) while preserving the underlying meaning. |
|  | Abbreviation Expansion/Contraction (T–Abbr) | Before: “right arm”;<br>After: “R arm” | Edits that expand, introduce, or standardize abbreviations and acronyms. |
| Language and Style | Empathy / rapport / politeness (S–Emp) | Before: *(none)*;<br>After: “It was a pleasure to see Ms. [NAME]” | Edits that change interpersonal tone (rapport, empathy, politeness, patient-centered phrasing) without changing core clinical content. |
|  | Certainty / hedging (S–Cert) | Before: “due to”;<br>After: “likely due to” | Edits that increase or decrease expressed certainty, probability, attribution, or diagnostic commitment (e.g., hedging, qualifiers, speculative language). |

**Table 2.**
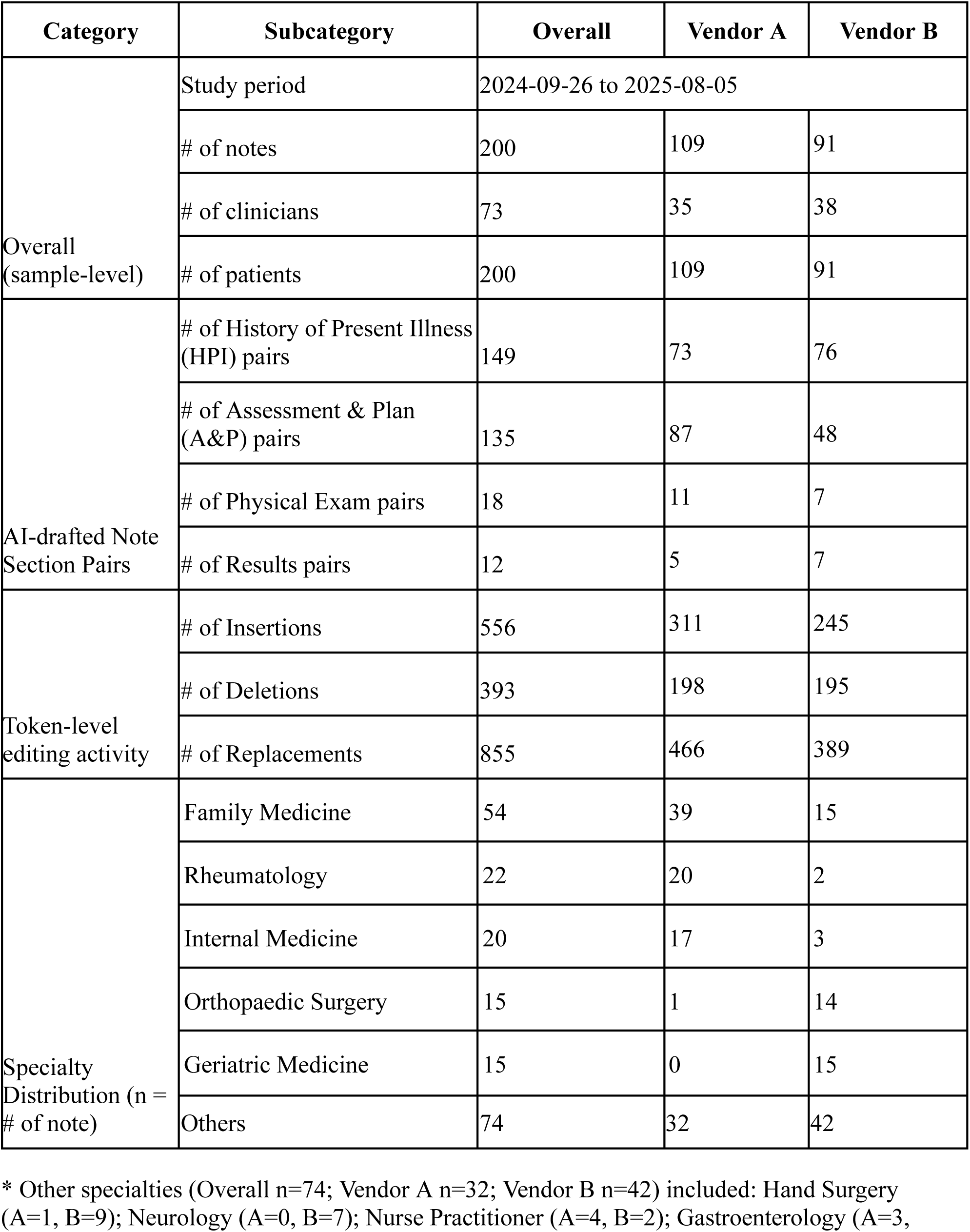
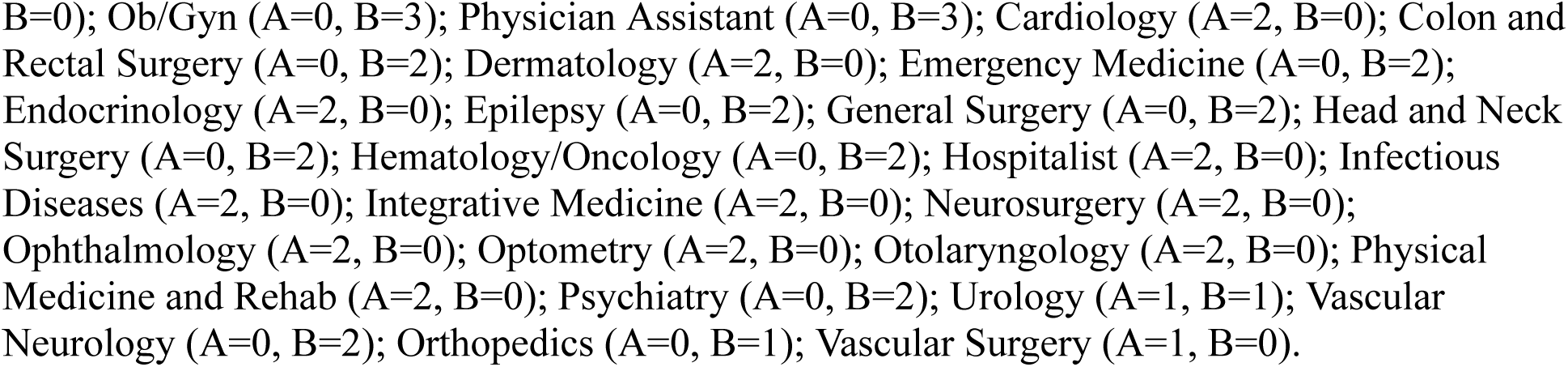
Characteristics for paired ambient AI drafts and clinician-finalized notes.

### Distribution of Qualitative Edit Codes

We coded 713 edit units extracted from 314 AI draft–final note section pairs. Vendor A contributed 373 edit units and Vendor B contributed 340 edit units. After an initial training phase and two rounds of independent coding with consensus discussions, interrater reliability across eight coders reached a moderate level of agreement (overall Cohen’s κ = 0.568), which is consistent with multi-label coding of heterogeneous clinical edits; disagreements were resolved through adjudication and consensus discussions. Because a single edit unit could receive multiple codes, results are reported as the frequency of edit units containing each code; percentages therefore do not sum to 100 percent. Table 3 summarizes the distribution of edit codes overall and by vendor.

**Table 3.** Distribution of qualitative edit codes overall and by vendor.

| **Code** | **Vendor A, n (%)** | **Vendor B, n (%)** | **Total, n (%)** |
| --- | --- | --- | --- |
| E-Test | 160 (42.9%) | 125 (36.8%) | 285 (40.0%) |
| E-Sym | 121 (32.4%) | 95 (27.9%) | 216 (30.3%) |
| E-Med | 120 (32.2%) | 75 (22.1%) | 195 (27.3%) |
| E-Dx | 111 (29.8%) | 74 (21.8%) | 185 (25.9%) |
| E-Hx | 73 (19.6%) | 62 (18.2%) | 135 (18.9%) |
| E-Soc | 60 (16.1%) | 67 (19.7%) | 127 (17.8%) |
| E-ID | 37 (9.9%) | 26 (7.6%) | 63 (8.8%) |
| T-Lay | 25 (6.7%) | 26 (7.6%) | 51 (7.2%) |
| T-Abbr | 22 (5.9%) | 10 (2.9%) | 32 (4.5%) |
| S-Cert | 17 (4.6%) | 12 (3.5%) | 29 (4.1%) |
| S-Emp | 11 (2.9%) | 11 (3.2%) | 22 (3.1%) |

\* Percentages are calculated using the total number of coded edit units within each subgroup as the denominator (Vendor A: N=373; Vendor B: N=340; Overall: N=713). Because an edit unit can receive multiple codes, counts may sum to >100%.

The most common edit types involved clinical content. Procedures, tests, and lab orders edits (E–Test) appeared in 285 edit units (40.0%). A common pattern was clarifying order-specific metadata, including the test type, the intended location or service, and the timing or status of the order when that detail affected interpretability. This was followed by symptom changes (E–Sym) in 216 (30.3%). For symptom edits, clinicians commonly refined the symptom description to improve clinical specificity. This included adjusting the symptom label, and specifying severity or pattern when the draft was vague. Clinicians also removed symptom descriptions that were not part of the patient’s current presentation or that appeared to be carried over from prior context without supporting evidence from the encounter. Medication modifications (E–Med) appeared in 195 (27.3%), and edits most often made the regimen actionable by adding or correcting dose, frequency, route, or administration details. Clinicians also standardized medication nomenclature, such as correcting the drug name, aligning the term with common clinical usage, or clarifying which formulation was intended. A smaller but recurring pattern was medication reconciliation, where clinicians removed a medication that was not relevant to the current plan or clarified whether the patient was currently taking it. Diagnosis refinements (E–Dx) appeared in 185 (25.9%). Clinicians frequently revised the diagnostic framing by narrowing a broad label to a more specific condition, aligning the diagnosis term with the symptom description, or clarifying whether the diagnosis was established versus under evaluation. Edits involving past medical history (18.9%) and social context information (17.8%) were also frequent, whereas terminology and wording edits such as jargon versus lay (7.2%) and abbreviation changes (4.5%) and language and style edits such as certainty or hedging (4.1%) and empathy and rapport (3.1%) were less common.

### Thematic Synthesis of Clinician Edits

Editing activity varied by note section, with the highest concentration in narrative sections such as HPI and Assessment and Plan and fewer edits in Physical Exam and Results. Vendor-stratified summaries showed broadly similar code distributions, and all five themes were observed in notes from both vendors. Given the qualitative aim and descriptive nature of vendor stratification, we present themes pooled across vendors for interpretability. Representative before/after examples are shown in Table 4.

**Table 4.**
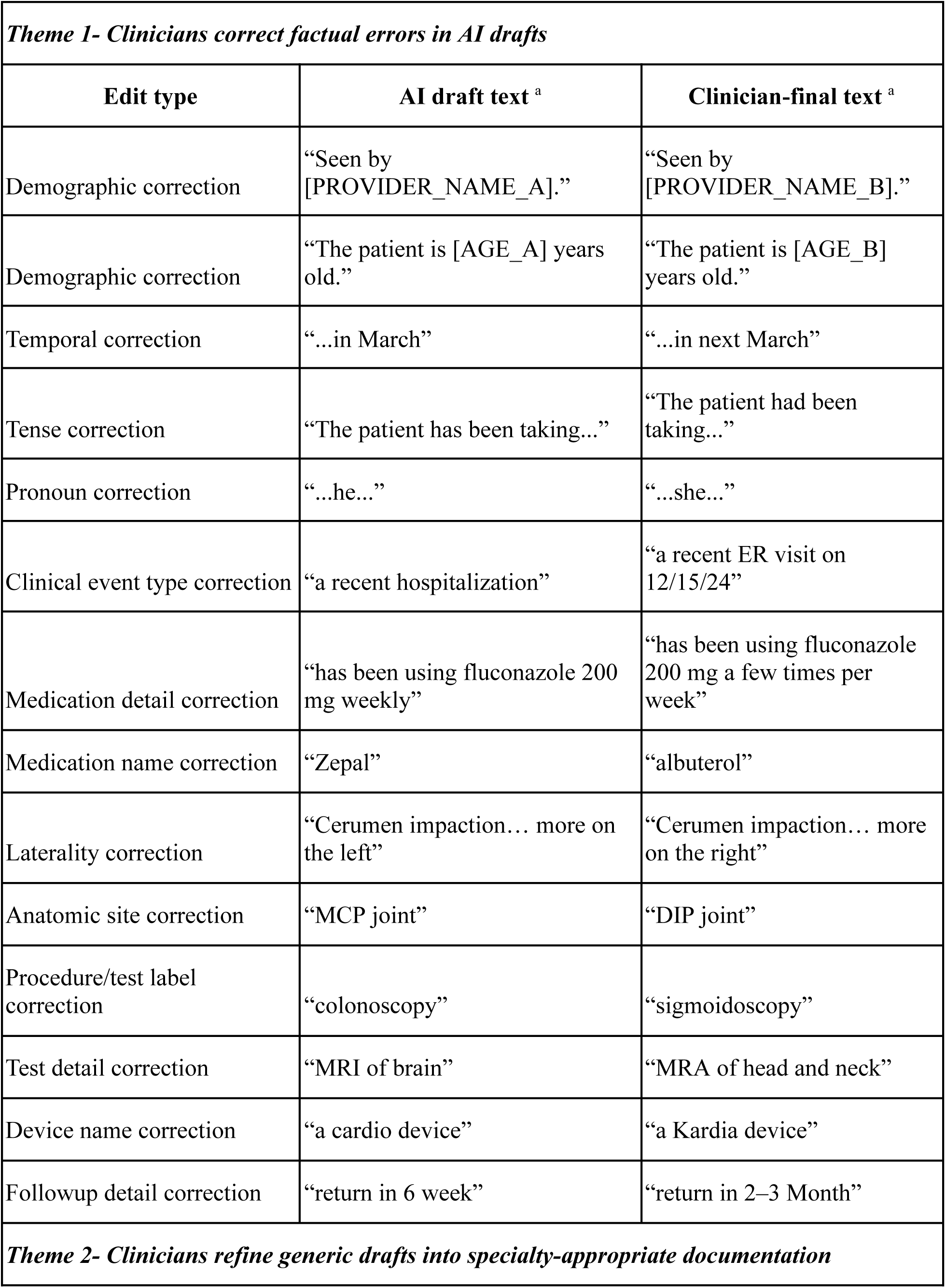
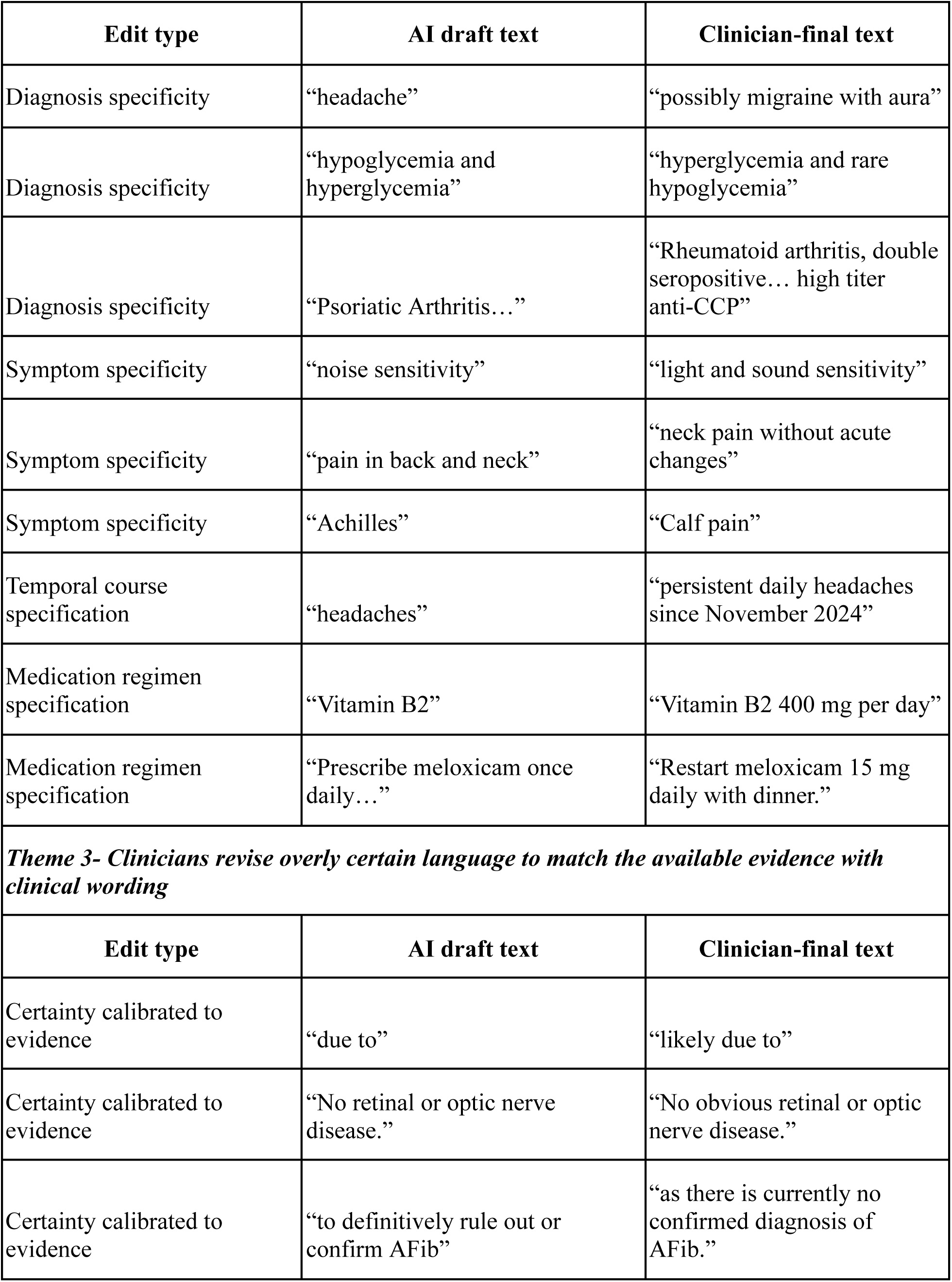
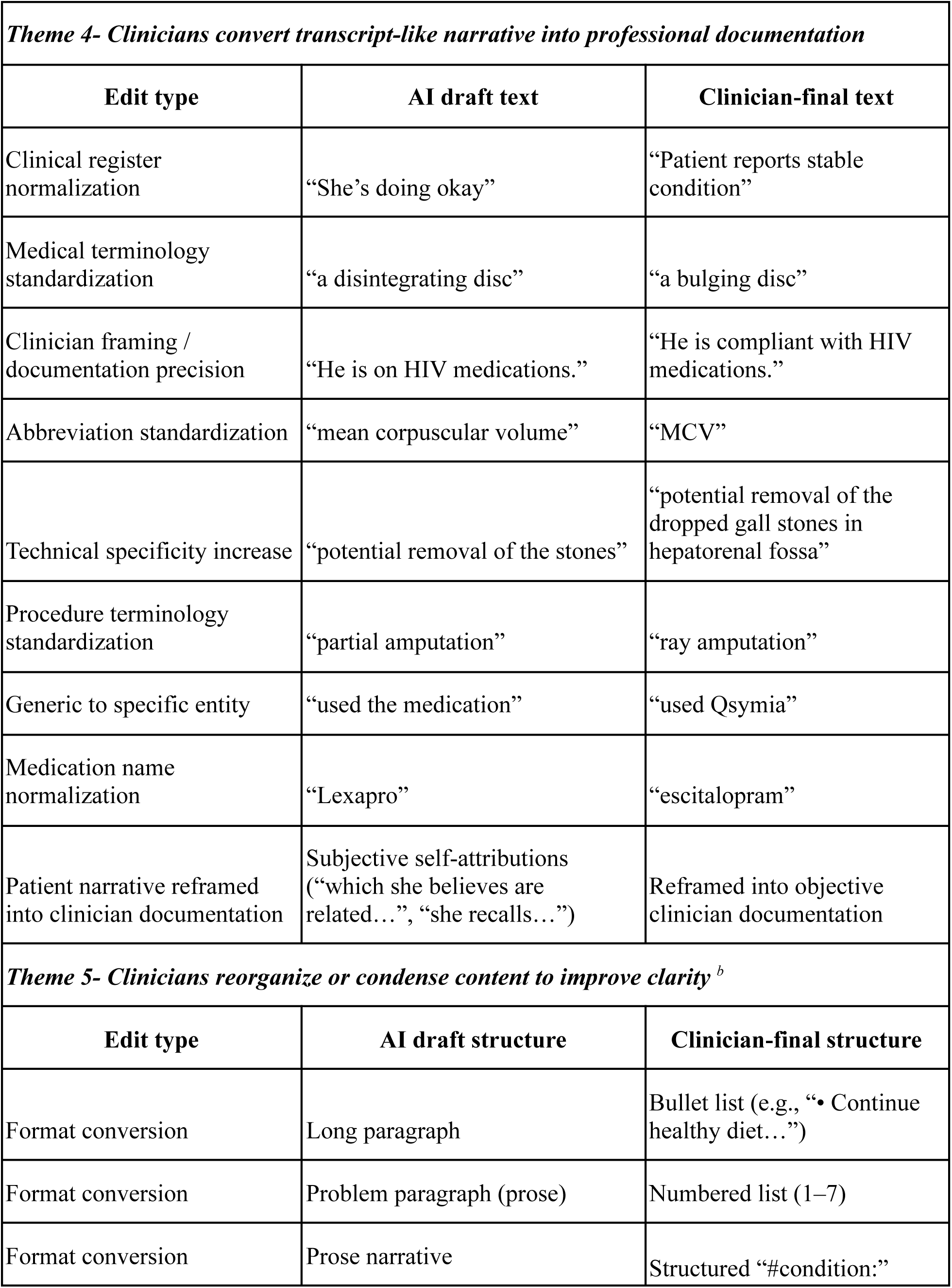
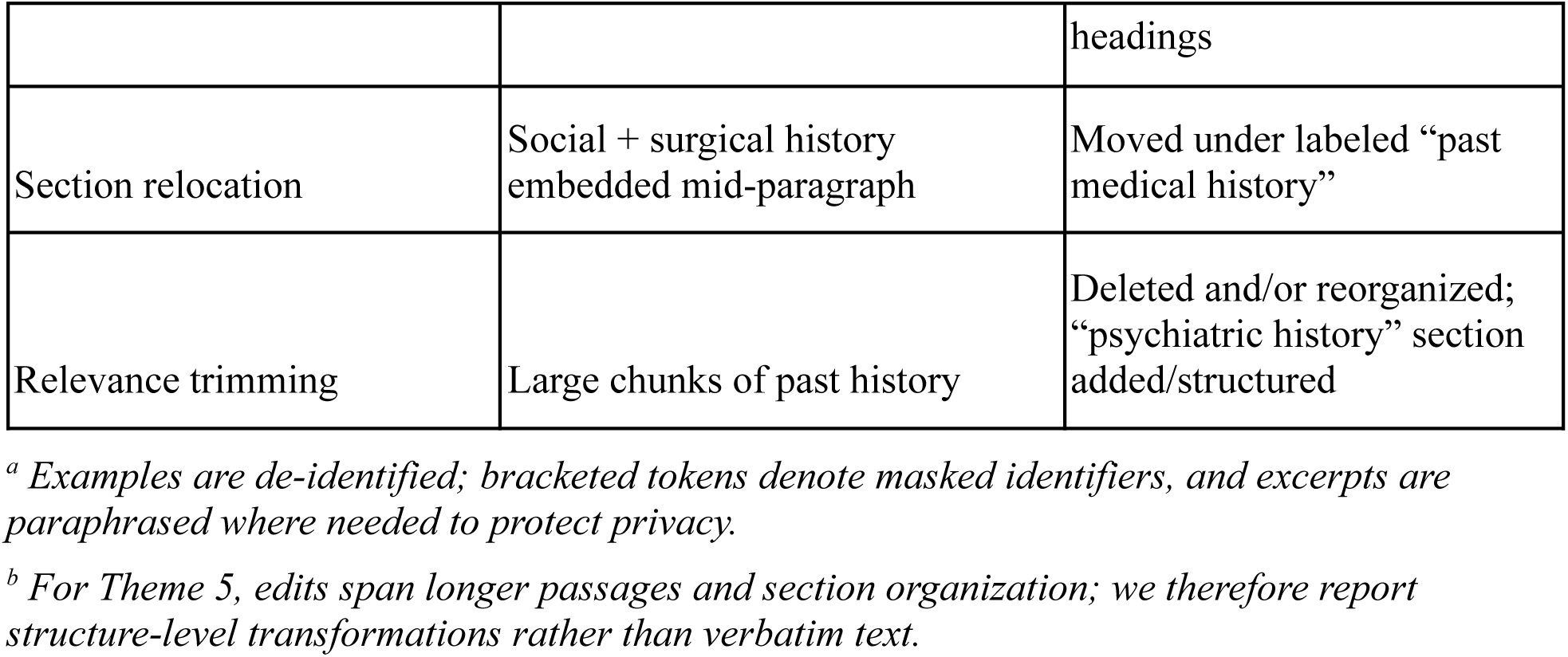
Representative examples of clinician edits to ambient AI–drafted notes by five qualitative themes.

#### Theme 1- Clinicians correct factual errors in AI drafts

These edits corrected misspelled names and pronouns introduced during transcription and fixed inaccurate time-stamp information. They also refined event descriptions by updating prior medical history details, such as changing a hospitalization to a more accurate emergency department visit, and by correcting anatomic and severity information (e.g., laterality, joint location, stenosis severity). Clinicians further corrected medication and test/procedure details, including drug names and dosing frequency, sedation type, and imaging modality. These edits improved the accuracy of the clinical note in aspects, including medical history, demographics, and treatments.

#### Theme 2- Clinicians refine generic drafts into specialty-appropriate documentation

Clinicians added clinical details commonly required for specialty documentation, such as symptom onset and trajectory, medication regimen specifics (dose, schedule, and whether use was as needed), and test or treatment context that shaped the plan. In neurology/epilepsy, edits emphasized temporal progression and neuroanatomical localization, reframing brief summaries into problem-oriented assessments that track symptom course, differentiate neurologic problems, and document medication intervals or titration. For example, a neurology progress note was expanded by adding detailed trauma history, symptom characterization across neurologic problems, and rewriting the assessment into a specialized, multi-layered plan. In procedure-oriented specialties such as hand surgery and orthopaedics, edits replaced general descriptions with structured, objective findings and procedure- or imaging-specific terminology, making decisions explicit through digit-level exam metrics or dated radiology findings. A hand surgery note was refined by replacing general symptom and exam language with digit-specific, quantified findings (e.g., finger-level localization, monofilament scores) and by stating clear procedural intent, such as proceeding with right index and ring trigger finger release. In cardiology, clinicians expanded symptom documentation beyond brief general phrasing by specifying exertional context and exercise tolerance, linking dyspnea, chest symptoms, or palpitations to what the patient can do and under what conditions, so the note reflected cardiovascular-style functional assessment rather than a generic symptom checklist.

#### Theme 3- Clinicians revise overly certain language to match the available evidence with clinical wording

Across notes, clinicians often replaced definitive causal or diagnostic wording with more appropriately qualified language when the documentation did not support a firm conclusion, such as changing “due to” to “likely due to” and revising “no retinal or optic nerve disease” to “no obvious retinal or optic nerve disease.” They also clarified when a condition was being evaluated rather than established, for example, by reframing plans aiming to “definitively rule out or confirm AFib” as monitoring in the setting of “no confirmed diagnosis of AFib.” Similarly, diagnostic labels were adjusted to match test evidence, such as shifting a stated dermatomyositis diagnosis to a suspected drug eruption after a negative myositis panel.

#### Theme 4- Clinicians normalize conversational narratives into standardized chart language

Clinicians frequently edited transcript-like language to align the note with clinician-facing documentation standards by removing or reframing patient-attributed causal explanations and speculative risk phrasing, and by replacing them with objective, clinically interpretable statements. For example, subjective clauses such as “which she believes are related to her thyroid condition” or “she recalls an instance when…” were removed or rewritten as verifiable clinical information, such as the current levothyroxine dose or the patient’s expressed concern about thyroid level fluctuations. More broadly, conversational and vague phrasing was rewritten into conventional chart language and medical terminology, such as changing “she’s doing okay” to “patient reports stable condition” and replacing lay descriptions like “a disintegrating disc” with “a bulging disc”.

Edits also standardized abbreviations and shorthand, for example, using “MCV” and inserting routine clinical abbreviations such as “EGD”, “abnl”, and “CIN II”. Clinicians increased technical specificity by naming the exact device, procedure, or structure, such as revising “partial amputation” to “ray amputation,” and specifying the ECU tendon. Medication references were similarly clarified by replacing generic wording with the specific drug name, such as “used the medication” to “used Qsymia” and “Lexapro” to “escitalopram,” with accompanying refinements to dosing or decision language.

#### Theme 5- Clinicians reorganize, relocate, and condense AI drafts

Clinicians frequently restructure AI-generated drafts to make the note easier to navigate and use, reorganizing content to improve scanability and clarity of clinical plans while trimming material that did not match the visit. Dense narrative was often converted into structured formats, for example, with long paragraphs rewritten as bullet lists, problem narratives turned into numbered plans, and free text reorganized under problem-oriented headings. Clinicians also relocated information to more appropriate sections, such as moving social or surgical history into labeled past history fields and separating imaging descriptions into a dedicated radiology-style section with dated, objective findings. Edits also reflected the process of relevance triage where duplicative or boilerplate text was consolidated, and content that appeared misaligned with the clinical context was removed, including passages on specialty-irrelevant medication management.

## Discussion

Clinicians substantially modify AI-generated drafts, and those edits follow repeated patterns that align with the five themes in our qualitative analysis, which point to specific opportunities for improvement in ambient AI documentation tools and their implementation.

Clinicians often correct inaccuracies or hallucinations, especially in discrete details that can change clinical meaning, such as names, medication dosages, and test or procedure attributes. Even when the overall narrative in AI drafts was broadly accurate, these errors are difficult for downstream readers to detect and can propagate if copied forward. This finding suggests that improving factual reliability of ambient AI tools should be a top priority, paired with workflow features that help clinicians quickly verify the fields that most often need to be double-checked. [21] At present, most ambient AI tools do not directly access the patient’s chart, so certain errors, such as names or other identifiers, cannot be automatically cross-checked even when the correct information already exists in the EHR. If chart integration becomes feasible, many of these errors could be corrected upstream. In the meantime, for institutions integrating ambient AI into routine practice, training for clinicians should go beyond a generic requirement to “review the draft” and instead teach a consistent verification routine that prioritizes the most error-prone details. [22,23]

Clinicians tailoring AI drafts to be more specialty-specific suggest that a single, generic note structure and style is often insufficient for specialty documentation needs. An actionable response is to support specialty-tuned drafting behavior through templates and prompts that reliably elicit the details that matter for a given discipline, rather than broadly relying on the subjective–objective–assessment–plan (SOAP) structured format. [24–26] For institutions, this implies that ambient AI adoption should be paired with specialty-level configuration and evaluation aligned with each specialty’s documentation norms and clinical priorities. [27]

Overly certain language in ambient AI drafts can outpace the available clinical evidence, creating extra work for clinicians and increasing the risk of carrying forward unverified conclusions. Beyond clinical risk, this pattern may raise medicolegal concerns if AI-generated phrasing systematically nudges notes toward stronger diagnostic commitments than the encounter supports. [28] In our data, clinicians spent substantial effort dialing back overly certain AI language. Actionably, AI developers can in response design models that default to appropriately qualified language and conventional diagnostic framing, and that avoid stating conclusions more strongly than the note supports. [29] At the practice level, this theme points to a clear focus for governance and education, reinforcing a consistent review habit around diagnostic uncertainty and ensuring the system’s outputs do not nudge documentation toward over-commitment when diagnoses are not yet established.

Clinicians often rewrote transcript-like text into a usable clinical record, shifting conversational phrasing into objective, clinically interpretable documentation. This mismatch may be amplified by how the system structures conversational content into the Subjective section (SOAP). [24] In our data, clinicians frequently condensed and reorganized that section, suggesting potential misalignment between how the draft is assembled and how clinicians typically write and review notes. Ambient AI developers can strengthen attribution-aware writing that keeps a clear boundary between patient reports and clinician assessment without adding interpretation that the note does not support. Terminology normalization can also help increase precision without causing note bloat. [30–33] For clinicians, this theme is a reminder that review is not only error correction; it is also ensuring the note reads as a professional record that others can quickly interpret and act on.

Recurring patterns of note restructuring and condensing suggest that draft organization is a major determinant of whether ambient notes are usable at the point of care. For example, ambient capture can include social history and social determinants of health (SDOH) information that is clinically relevant because it can shape access to care, home and community support, and adherence. However, in the context of certain encounters, some of these elements may be secondary to the immediate diagnostic and clinical decision making, particularly when captured with high granularity (e.g., transportation needs narratives). [34] Clinicians often shortened or removed these details when they did not inform the assessment or plan. These edits reflect a familiar tradeoff in real-world documentation. As notes get longer, they become harder to review efficiently, and important information can be harder to find. [35,36] An actionable implication is to prioritize tool behaviors that produce more structured drafts, route content into predictable sections, and give clinicians simple ways to control verbosity without losing clinically important context. At the organizational level, this supports setting expectations for note structure and routinely monitoring note length and usability as part of ambient AI governance.

This study has several limitations. Our qualitative analysis drew on the AI draft–final note section pairs from 200 notes, and some specialties were represented by only a small number of cases, so we may not fully capture specialty-specific nuances. The dataset also comes from a single health system and a single deployment context, which may limit how well these patterns generalize to other institutions, care settings, or ambient AI products. Vendor-stratified summaries are descriptive, and the study was not designed or powered for formal between-vendor comparisons. The modest qualitative sample and broad thematic categories might limit sensitivity to subtle vendor-specific differences. Interrater agreement was moderate (overall Cohen’s κ = 0.568). This is consistent with a multi-coder, multi-label codebook applied to heterogeneous notes spanning diverse specialties and patient contexts, where code boundaries can overlap for clinically adjacent edits. We mitigated this through iterative calibration, adjudication, and consensus discussions, and we generated themes based on convergent patterns across codes and examples rather than any single coder’s label. [37] We only analyzed text changes without the source audio, user interface cues, or clinicians’ stated intent. Therefore, cannot determine why a given edit was made or whether it reflects individual style, local documentation norms, or risk tolerance. Finally, we did not assess the clinical correctness of the finalized notes or downstream impacts such as care outcomes, time burden, or corresponding change of billing codes.

Taken together, our findings translate routine clinician edits into concrete actionable insights for both tool improvement and real-world implementation. The patterns we observed suggest that ambient AI should be evaluated and tuned for specialty needs and clinician-to-clinician variation. Future work should incorporate targeted semi-structured interviews to clarify the rationales and workflow factors behind common edits and to validate our interpretation of these patterns. Future work should also investigate whether specific edit patterns are associated with continuity of care. For example, edits that clarify the problem list or tighten the assessment and plan may make notes more usable for downstream readers. Studies should also examine operational performance, including turnaround time, downstream review effort or follow-up work, and whether term-level changes affect billing or coding outcomes. Finally, future work should evaluate safety-critical signals, especially the correction of factual inaccuracies and the moderation of overly certain language when diagnoses remain uncertain.

## Conclusion

Our qualitative content analysis of clinician edits to AI drafts provides a comprehensive view of what clinicians changed and where improvement is needed. Clinicians corrected mistakes, replaced overly generic statements with specialty-appropriate detail, and tempered overly certain language to better match the available evidence using standard clinical wording. They also rewrote conversational, patient-attributed narrative into professional documentation and reorganized drafts into clearer, more structured notes. Ambient AI tools should be designed around how clinicians document in different specialties and should avoid wording that goes beyond the evidence in the encounter. Health systems can use recurring edit patterns to identify target implementation support and training that improve day-to-day documentation efficiency.

## CONTRIBUTORSHIP STATEMENT

Yawen Guo (YG) led the study, contributing to the study concept and design, data acquisition, data analysis and interpretation, and drafting of the manuscript. Di Hu (DH) contributed to notebook development and qualitative coding. Ziqi Yang (ZY), Brian Tran (BT), Jamie Lee (JL), Sitha Vallabhaneni (SV), Seungjun Kim (SK), and Rachael Zehrung (RZ) contributed to qualitative coding. Sairam Sutari (SS) provided data curation and administrative support. Steven Tam (ST), Emilie Chow (EC), Danielle Perret (DPe), and Deepti Pandita (DPa) provided clinical perspective and contributed to interpretation of findings. Kai Zheng (KZ) provided overall guidance and supervision throughout the project. All authors critically reviewed and revised the manuscript for important intellectual content and approved the final version.

## DATA AVAILABILITY STATEMENT

The underlying dataset contains protected health information (PHI) and is not publicly available.

